# Lifelong trajectories of modifiable vascular risk factors and relation to cerebral small vessel disease in the Framingham Heart Study

**DOI:** 10.64898/2026.01.16.26344309

**Authors:** Jose R. Romero, Adlin Pinheiro, Serkalem Demissie, Hugo J. Aparicio, Vasileios-Arsenios Lioutas, Antreas Charidimou, Alexa Beiser, Oluchi Ekenze, Jayandra J. Himali, Charles DeCarli, Sudha Seshadri, Shariq Mohammed

## Abstract

**Background:** Cerebral small vessel disease (CSVD) is strongly linked to stroke and dementia risk, frequently predating clinical events for years to decades. Preventive efforts focus on treatment of modifiable vascular risk factors (VRF), but the complex changes in VRF over long periods in relation to CSVD burden remain unclear. Thus, we aimed to characterize life-long VRF trajectories in community-dwelling individuals and relate them to CSVD burden.

**Methods:** Framingham Heart Study participants from the Original and Offspring cohorts with six or more repeated VRF assessments over their lifetime were eligible for the present study. Among those who underwent MRI imaging, CSVD burden was quantified by assigning one point each for cerebral microbleeds, covert infarcts, extensive white matter hyperintensities, cortical superficial siderosis, and high perivascular space burden (range: 0–5). VRF trajectories and trajectory-based clusters were then examined in relation to CSVD burden using functional regression analysis.

**Results:** Of 7,961 participants with longitudinal VRF measurements (mean baseline age 39.8 ± 10 years, 45% men), 1,625 underwent MRI imaging after exclusions for other neurological conditions. In models that used individual VRF trajectories as covariates, systolic and diastolic blood pressure, pulse pressure, cigarettes per day and triglycerides were significantly associated with CSVD burden later in life. In a complementary analysis, we clustered VRF trajectories and then used each participant’s cluster assignment (rather than the raw trajectories) as the covariate; cluster assignments diverged early in life for systolic blood pressure and pulse pressure and were significantly associated with CSVD burden.

**Conclusions:** Individuals following high risk lifetime patterns of vascular risk factors have higher CSVD burden later in life. These results highlight the importance of primordial and primary prevention with sustained risk factor management across the lifespan to mitigate CSVD burden, a strong determinant of stroke and dementia risk.

## Introduction

Traditional vascular risk factors (VRF) are strongly linked to increased risk of major cardiovascular events and remain as the main targets for prevention of cardiovascular disease, stroke and cognitive disorders. While several effective treatments exist for VRF, stroke remains a major public health concern, considered the 4^th^ most common cause of death in the US,^1^ with a prevalence projected to increase dramatically as the aging population and survival after stroke increase.^2^ Similarly, dementia is increasing exponentially with doubling of prevalence expected by year 2050.^3^ Thus, early identification of individuals at increased risk is imperative to implement preventive strategies at the earliest stages of disease.

Cerebral small vessel disease (CSVD) is one of the most common types of cerebrovascular disease increasing risk of stroke and dementia. Detection of CSVD in subclinical stages can be done years or even decades before clinical events manifest. In turn, VRF predate subclinical CSVD for years to decades and influence progression of CSVD burden once established as well as subsequent risk of clinical events. Thus, further detailed understanding of cumulative exposure to VRF and its relation to CSVD burden in subclinical stages presents an opportunity for early preventive interventions (i.e. primordial and primary) for subsequent stroke and dementia. One key challenge is understanding how vascular risk factors evolve over time and how these changes affect CSVD burden later in life.

In the prospective, longitudinal Framingham Heart Study, we leveraged repeated VRF measurements over a lifetime and CSVD markers measured on MRI to investigate how long-term risk factor patterns relate to CSVD burden later in life. This approach allows for a more comprehensive understanding of the sustained effects of vascular risk over time. Identifying distinct trajectory patterns and their associations with CSVD burden may offer key insights into the timing and nature of interventions needed to reduce CSVD-related risk.

## Methods

### Sample Selection

Original and Offspring cohort participants of the Framingham Heart Study attending more than six clinic examinations over the course of their participation were eligible for the present study. The Original cohort has been examined every two years for 32 exam cycles since 1948 while the Offspring cohort, now in its tenth examination cycle, has been examined every 4-7 years since 1971. Out of a total 10,333 Original and Offspring participants, 7,961 attended at least six clinic exams. A subset of participants also had brain MRI with 1,625 having multi-marker CSVD scores after excluding 46 participants for other neurological conditions that could affect brain MRI (Figure 1). When multiple MRIs were available, the most recent scan was selected.

**Figure 1:**
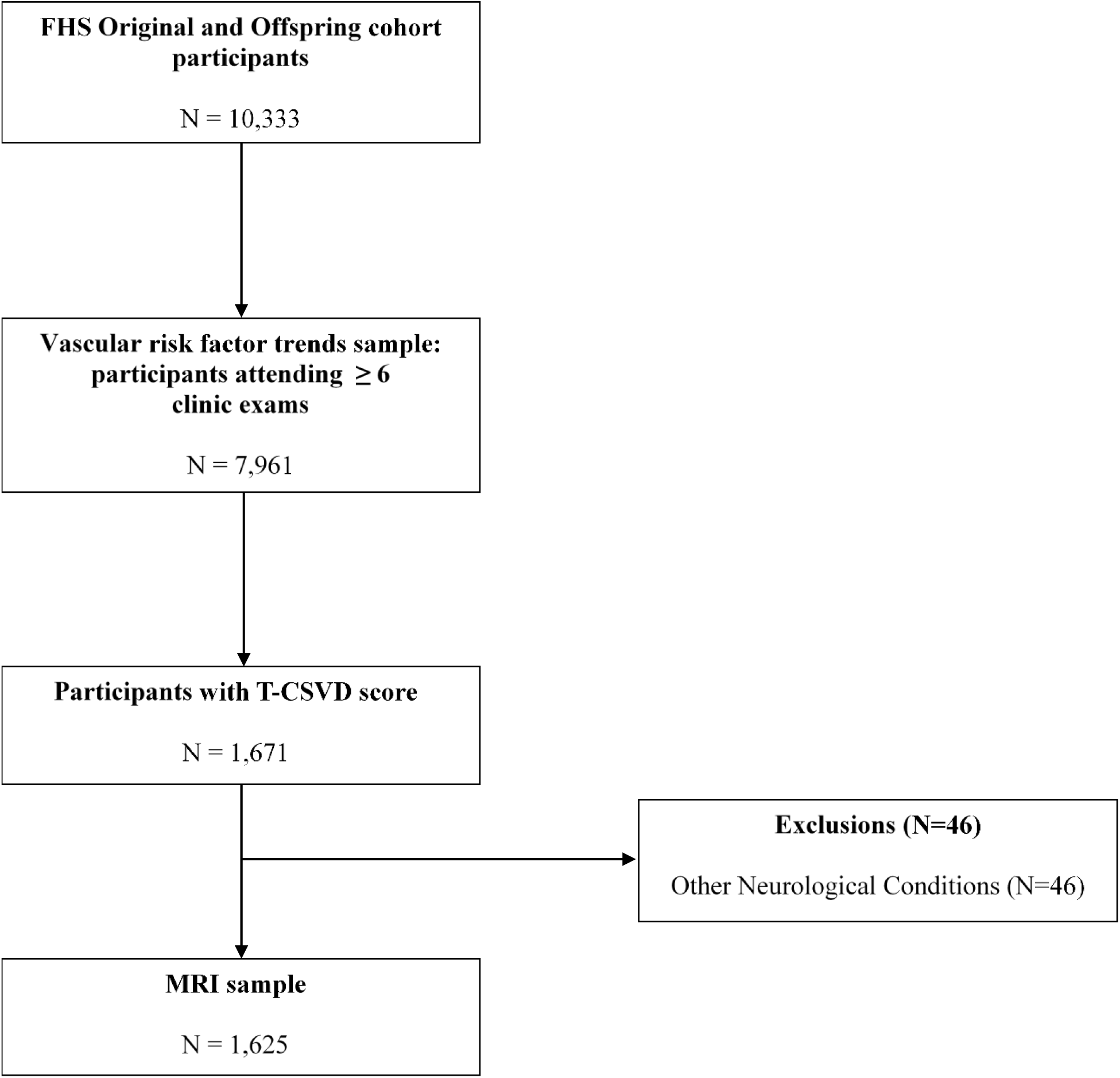
Sample selection flow chart.

### Vascular Risk Factors

Vascular risk factors were measured at the time of the clinic exams. Data from exams 1 through 32 (1948-2014) were used for the Original cohort while data from exams 1 through 9 (1971-2014) were used for the Offspring cohort. Risk factors included systolic blood pressure (SBP, mmHg), diastolic blood pressure (DBP, mmHg), pulse pressure, body mass index (BMI, kg/m^2^), number of cigarettes smoked per day, serum levels of triglycerides (mg/dL), high density lipoprotein (HDL, mg/dL), low-density lipoprotein (LDL, mg/dL), total cholesterol (mg/dL), and blood glucose (mg/dL). Details on the clinic exams used to obtain each risk factor for the sample are provided in Supplementary Tables S1 and S2.

Blood pressure was measured by a physician at each examination cycle with a mercury column sphygmomanometer and appropriate width-sized cuff. Participants were in a seated position, and measurements were done in the left arm and rounded to the nearest even number. The mean of two blood pressure measurements was taken as the value for each exam. Pulse pressure was determined as the difference between SBP and DBP.

Random blood glucose was measured in the Original cohort and fasting blood glucose was measured in the Offspring cohort, starting from the third clinic exam. LDL was considered only for the Offspring cohort since LDL was only measured in later exams for the Original cohort. Triglyceride levels were limited to a maximum of 400 mg/dL.

Number of cigarettes smoked per day and use of anti-hypertensive, lipid lowering and diabetes medication was self-reported. Medication use reported during a clinic exam was assumed to extend throughout the entire exam cycle, lasting two years for the Original cohort and four years for the Offspring cohort. Supplementary Tables S1 and S2 contain information on data availability for each exam cycle.

### Multi-marker CSVD score

Brain MRI have been acquired in FHS participants since 1999 using 1T, 1.5T or 3T scanners, with acquisition parameters, image processing previously published.^4^ Ratings for CSVD markers was blinded to all subject’s demographic and clinical characteristics, using published criteria by the Standards for Reporting Vascular Changes on Neuroimaging (STRIVE-2) consortium.^5^ The intra and inter-rater reliability for all CSVD markers ranged from good to excellent as previously reported.^6^

Because of heterogeneity in the presence and burden of single CSVD markers, we used a multi-marker CSVD score to capture the overall burden reflected by the best characterized CSVD markers. The score assigned one point to the presence of each of the following markers detected in the MRI: covert brain infarcts (CBI), cerebral microbleeds (CMB), extensive white matter hyperintensities (WMH), cortical superficial siderosis (cSS), and high burden of MRI visible perivascular spaces (PVS). Methods evaluating these markers in the FHS have previously been reported.^6^

CBIs were identified based on lesion size (≥3 mm), location, and imaging characteristics, including cerebrospinal fluid-like intensity on subtraction images (proton density-T2) and hyperintensity on T2-weighted images. CMBs were defined as rounded or ovoid hypointense lesions, less than 10mm in diameter, surrounded by brain parenchyma over at least half the circumference of the lesion, excluding CMB mimics. cSS was defined as a linear, gyriform low-intensity signal visible on T2*-weighted gradient-echo MRI. PVS were defined as linear or ovoid lesions less than 3 mm in diameter, exhibiting signal intensity similar to cerebrospinal fluid across all sequences and following the path of penetrating vessels. High burden of PVS was defined as Grade III-IV PVS (count > 20) in either the basal ganglia or centrum semiovale. WMH volume was calculated as a percentage of total intracranial volume. Extensive WMH was defined as a log-transformed WMH areas exceeding one standard deviation above the age-specific mean.

### Statistical Analysis

Descriptive statistics for clinical and demographic variables at the baseline exam of the included and excluded samples are presented using mean (SD) for continuous variables and counts (percent) for categorical variables. We estimated the mean longitudinal trajectory of each risk factor as a function of age along with corresponding 95% bootstrap confidence intervals (i.e., participants were aligned by age at the onset of each VRF trajectory); all VRFs were treated as continuous, including cigarettes per day (a count variable treated as continuous for analysis). Mean trajectories were also stratified by sex and cohort (supplemental figures 1 and 2).

Functional data analysis (FDA) was used to examine the lifetime trajectories of vascular risk factors across participants. For each risk factor assessed, participants were required to have at least six recorded measurements over the course of their participation in the study for inclusion. Risk factor values measured at all available clinic exams were plotted against age to construct lifetime trajectories for each individual. Functional principal components analysis (FPCA), a commonly used dimension-reduction technique in FDA, was employed to generate Euclidean representations of participant’s trajectories for each risk factor, treating these trajectories as mathematical functions. Similar to traditional PCA, FPCA captures the dominant patterns of variation in functional data. This approach allows for efficient summarization of trajectory data while preserving the key characteristics of individual patterns. We used FPCA scores in two distinct analyses. First, to identify associations of VRFs with CSVD, we conducted a functional regression analysis with FPCA scores as predictors and CSVD score as outcome.

Second, to enhance clinical interpretation, we grouped participants into VRF trajectory categories (e.g., low, moderate, high) by clustering the FPCA scores. Specifically, we employed the PACE (Principal Analysis by Conditional Expectation) algorithm for FPCA.^7^ PACE accommodates both irregularly observed and sparsely sampled data and does not require pre-smoothing of individual trajectories. PACE is particularly suited for longitudinal datasets where the timing and number of measurements vary across individuals. The algorithm estimates the mean function and covariance structure of the functional data using local smoothing techniques. By leveraging these estimates, PACE reconstructs individual trajectories and identifies principal components that represent dominant modes of variation in the data. In this study, PACE was used to generate functional principal component scores to summarize the variability in lifetime trajectories across participants for use in statistical analysis. The number of principal components to include in analyses was chosen to explain 99% of the variability in the trajectories.

The FPC scores based on the entire sample were then used as predictors in ordinal logistic regression models to evaluate associations with multi-marker CSVD score among participants who had undergone an MRI and had CSVD score ratings. This functional regression approach quantifies associations with the VRF features (i.e., FPCA scores). A schematic representation of the analyses is presented in Figure 2. The proportional odds assumption was assessed using the score test. Due to the small number of participants with score≥2, we combined these participants into one group, creating a new score with the following categories: 0 (no CSVD markers), 1 (one CSVD marker), and 2+ (two or more CSVD markers). This grouping is supported by increasing risk of incident stroke and dementia observed with increasing number of single CSVD markers present.^6,8^

**Figure 2:**
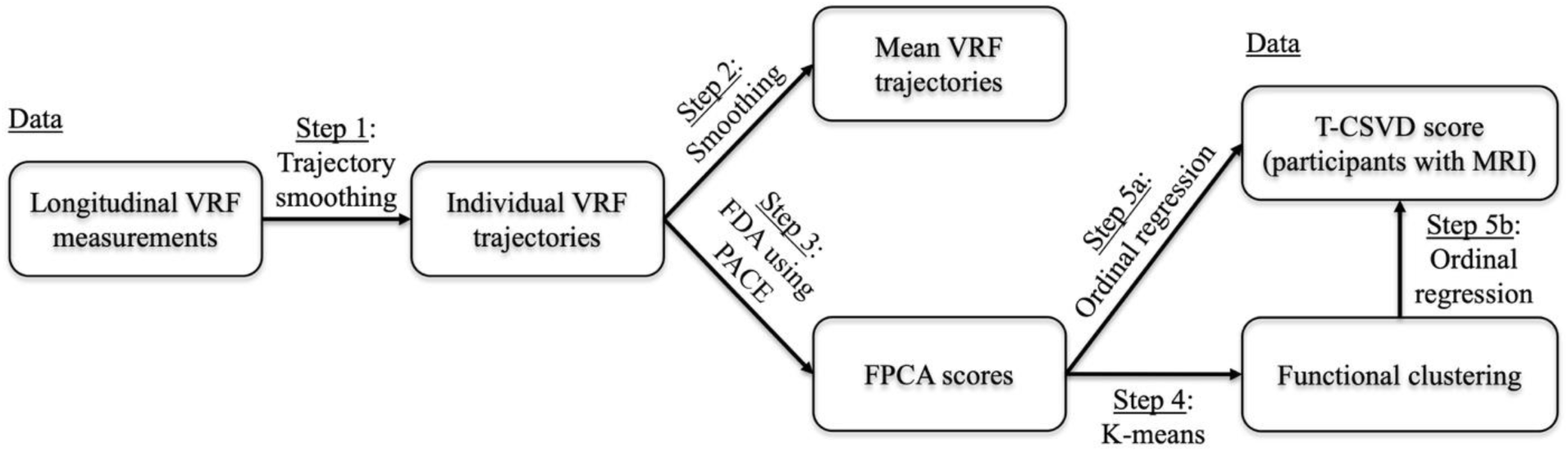
Analytic workflow for vascular risk factor (VRF) trajectories and their association with cerebral small vessel disease (CSVD). All analyses were conducted separately for each vascular risk factor (VRF). Data prospectively measured VRF at every exam cycle from study entry, and CSVD burden was summarized among participants with MRI using a multi-marker score (categorized as 0, 1, or ≥2 markers; CSVD). The workflow comprised: <u>Step 1</u>: Alignment of individual VRF trajectories by age. <u>Step 2</u>: estimation of the mean VRF trajectory via smoothing. <u>Step 3</u>: Functional principal component (FPC) analysis using PACE to derive subject-specific FPC scores while accommodating irregularly spaced and sparsely observed longitudinal measurements, retaining components that cumulatively capture most variance. <u>Step 4</u>: k-means clustering of FPC scores to identify groups reflecting distinct long-term exposure patterns. <u>Step 5</u>: Ordinal multivariable logistic regression testing the global association of (<u>5a</u>) FPC scores and (<u>5b</u>) cluster membership with the multi-marker CSVD score as the outcome.

The first model adjusted for age at MRI, sex, and FHS cohort. A second model additionally adjusted for years of use of anti-hypertensive, lipid lowering, and diabetes medications. Global hypothesis tests were conducted to determine the significance of all FPC scores of a particular risk factor. Odds ratios and 95% confidence intervals are presented for a one standard deviation change in the principal component score. Note that the CSVD score was assessed at the date of the most recent MRI, and FPCA scores were computed using all available clinic exams to summarize each participant’s lifetime trajectory. The primary analyses therefore related VRF-based FPCA scores to CSVD burden measured at MRI, and should be interpreted as associations with long-term patterns rather than strictly pre-MRI exposures.

In the secondary analysis, k-means clustering was performed on the FPC scores for each risk factor in the entire sample, assigning participants into groups based on their longitudinal trajectories. We employed functional clustering to identify empirical groupings based on the observed similarity of VRF trajectories, without imposing the parametric assumptions typically required by latent growth models. Clustering on the FPCA scores leverages the same trajectory information used in the regression models yielding clinically interpretable VRF trajectory-based groups. Cluster sizes of three or four were selected for each risk factor based on clinician judgment and expertise, informed by elbow and silhouette plots. Four clusters were selected for trajectories of cigarettes per day, blood glucose, and triglycerides; three were selected for the remaining risk factors. Cluster labels were then assigned based on the trajectory peaks of each risk factor. For all risk factors (except HDL) with three clusters, the cluster with the highest peak was labeled as ‘high risk,’ the cluster with the lowest peak was labeled as ‘low risk,’ and the intermediate cluster as ‘moderate risk.’ For risk factors with four clusters, the cluster with the highest peak was labeled as ‘very high risk”. For HDL, the cluster with the lowest peak was labeled as “high risk” and the cluster with the highest peak was labeled “low risk”.

Similar to the primary analysis, cluster assignment (informed by the entire sample) was then used as a predictor in ordinal logistic regression models to evaluate associations with multi-marker CSVD score adjusting for age at MRI, sex, and cohort. A secondary model also adjusted for years of medication use.

For each VRF, we fit an ordinal logistic regression that included all retained FPC scores for that VRF (typically 3–5) simultaneously, along with covariates. Because FPC scores are orthogonal by construction, simultaneous inclusion does not introduce multicollinearity. A global hypothesis test would test the overall association of each VRF with CSVD-score. Global p-values across VRF models were adjusted for multiple testing using the Benjamini–Hochberg false discovery rate procedure, and an adjusted p-value < 0.05 was considered statistically significant. Within each VRF model, we report odds ratios (ORs) with 95% confidence intervals; accordingly, we did not perform multiple-testing correction at the coefficient level because we do not use p-values to determine significance. R package *fdapace* was used to calculate the functional principal components scores. SAS version 9.4 (SAS Institute, Cary, NC) was used for the remaining analyses.

## Results

Table 1 contains the baseline characteristics for the overall sample at the time of the first clinic exam. Mean age at the baseline exam was 39.8 years (SD: 10.0), 45% were male, and 56% were from the Original cohort. Among participants who had undergone an MRI later in life, average age at MRI was 72.8 (SD: 9.4), 45% were male, and 91% came from the Offspring cohort (Table 2).

**Table 1:** Baseline descriptive statistics for vascular risk factor trajectory sample.

| **Baseline Characteristics at baseline exam** | **Included** |  | **Excluded** |  |
| --- | --- | --- | --- | --- |
|  | **N** | **Mean (SD)** | **N** | **Mean (SD)** |
| Age, years, mean (SD) | 7961 | 39.8 (10.0) | 2372 | 41.6 (11.3) |
| Male, n (%) | 7961 | 3566 (45) | 2372 | 1253 (53) |
| FHS Cohort, n (%) | 7961 |  | 2372 |  |
| Original |  | 4434 (56) |  | 775 (33) |
| Offspring |  | 3527 (44) |  | 1597 (67) |
| Systolic blood pressure, mmHg, mean (SD) | 7961 | 127.7 (19.6) | 2368 | 131.7 (23.4) |
| Diastolic blood pressure, mmHg, mean (SD) | 7961 | 81.5 (11.7) | 2368 | 83.2 (13.4) |
| Pulse pressure, mean (SD) | 7961 | 46.3 (11.9) | 2368 | 48.4 (14.2) |
| Body mass index, kg/m<sup>2</sup>, mean (SD) | 6579 | 25.0 (4.0) | 2368 | 25.7 (4.7) |
| Cigarettes per day, mean (SD) | 7485 | 8.5 (12.0) | 2367 | 13.0 (14.7) |
| Low density lipoprotein*, md/dL, mean (SD) | 3327 | 124.9 (35.4) | 1529 | 133.5 (40.2) |
| High density lipoprotein, md/dL, mean (SD) | 3841 | 52.5 (16.0) | 1564 | 50.2 (15.9) |
| Total cholesterol, md/dL, mean (SD) | 7581 | 211.4 (44.4) | 2119 | 210.9 (46.2) |
| Random blood glucose<sup>†</sup>, md/dL, mean (SD) | 3885 | 80.9 (18.2) | 649 | 84.6 (30.1) |
| Fasting blood glucose<sup>†</sup>, md/dL, mean (SD) | 2414 | 91.9 (15.6) | 566 | 102.1 (34.3) |
| Triglycerides, md/dL, mean (SD) | 3473 | 92.8 (63.4) | 1569 | 115.5 (114.4) |

For the included sample, baseline values are the first values included in the individual trajectories. For the excluded sample, baseline values are measured at exam 1 for systolic and diastolic blood pressure, pulse pressure, body mass index, cigarettes per day, low density lipoprotein, total cholesterol, high-density lipoprotein (Offspring), triglycerides (Offspring) and random blood glucose; exam 3 for fasting blood glucose, exam 7 for triglycerides (Original), and exam 15 for high-density lipoprotein (Offspring).
\*Offspring only
<sup>†</sup> Fasting blood glucose was measured in the Offspring cohort, random blood glucose in the Original cohort.

**Table 2:** Baseline descriptive statistics of the MRI sample.

| **Baseline Characteristics** | **Included**<br>**(N=1,625)** | **Excluded**<br>**(N=46)** |
| --- | --- | --- |
| Age, years, mean (SD) | 33.3 (8.9) | 32.3 (8.2) |
| Male, n (%) | 730 (45) | 20 (43) |
| FHS cohort, n (%) |  |  |
| Original | 142 (9) | 4 (9) |
| Offspring | 1483 (91) | 42 (91) |
| Systolic blood pressure, mmHg, mean (SD) | 118.3 (13.2) | 120.4 (15.8) |
| Diastolic blood pressure, mmHg, mean (SD) | 76.4 (9.4) | 78.1 (10.7) |
| Pulse pressure, mean (SD) | 41.9 (8.6) | 42.3 (9.3) |
| Body mass index, kg/m<sup>2</sup>, mean (SD) | 24.2 (3.9) | 25 (4.3) |
| Cigarettes per day, mean (SD) | 6.9 (11.3) | 6.3 (9.2) |
| Low density lipoprotein, md/dL, mean (SD) | 120.6 (34.3) | 119.2 (32.1) |
| High density lipoprotein, md/dL, mean (SD) | 52.9 (16) | 53.1 (17.5) |
| Total cholesterol, md/dL, mean (SD) | 192.7 (37) | 191.5 (38.6) |
| Blood glucose*, md/dL, mean (SD); N = 142 | 78.8 (13.3) | ** |
| Fasting blood glucose*, md/dL, mean (SD); N=1483 | 91.4 (15.2) | 94.9 (16.1) |
| Triglycerides, md/dL, mean (SD) | 91.2 (59.4) | 93.5 (70.8) |
| *MRI Markers* |  |  |
| Age at MRI, years, mean (SD) | 72.8 (9.4) | 72.1 (9.5) |
| Multi-marker CSVD Score, n (%) |  |  |
| 0 | 761 (47) | 13 (28) |
| 1 | 506 (31) | 11 (24) |
| 2+ | 358 (22) | 22 (48) |
| Covert brain infarct, n (%) | 212 (13) | 19 (41) |
| Cerebral microbleed, n (%) | 194 (12) | 9 (20) |
| High PVS burden, n (%) | 653 (40) | 19 (41) |
| Extensive white matter hyperintensities, n (%) | 272 (17) | 20 (43) |
| Cortical superficial siderosis, n (%) | 2 (0) | 0 |
| Dementia, n (%) | 73 (4) | 4 (9) |
| History of stroke, n (%) | 71 (4) | 2 (4) |

FHS = Framingham Heart Study; MRI = magnetic resonance imaging.
\*Fasting blood glucose was measured in the Offspring cohort, random blood glucose in the Original cohort
\*\*Insufficient data.

### Vascular risk factor trajectories

Figure 3 contains mean trajectories and 95% confidence bands for the entire sample by risk factor. Supplemental Figures S1 and S2 contain mean trajectories stratified by FHS cohort and sex. For all risk factors, confidence bands at older ages (> 80 years) were wider, reflecting the small number of surviving participants at those ages.

**Figure 3:**
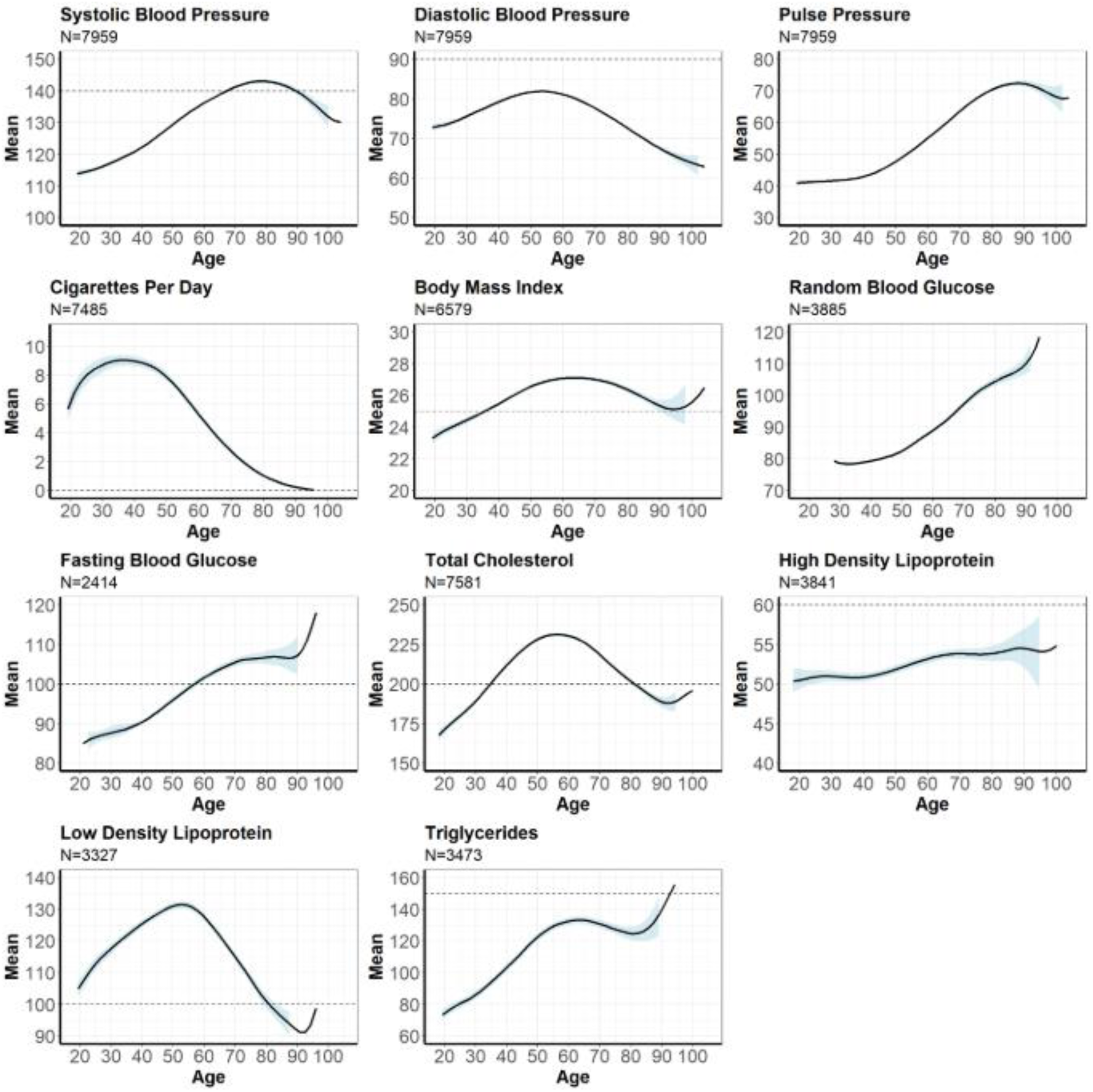
Mean Vascular Risk Factor Trajectories and 95% Confidence Bands.

Mean SBP steadily increased throughout the life course, starting at 115 mmHg at age 20 and peaking at age 80 at 145 mmHg before decreasing. Similar trends were observed with DBP, which peaked at age 55. The number of cigarettes per day increased from six in early life to nine at 40 before gradually decreasing to zero in late life. Mean BMI steadily increased from early adulthood to age 60, from 23 to 27 kg/m^3^, before decreasing to 25 kg/m^3^ at age 90. Mean fasting and non-fasting blood glucose both increased over the life course without decline. Mean HDL observed steady increases throughout the life course, starting at 50 mm/dL in early life to just under 55 mg/dL at age 90. Mean total cholesterol levels observed a parabolic trend, increasing from 170 mg/dL in early life to 230 mg/dl at age 60 before decreasing to 200 mg/dL at age 80. Mean triglycerides increased from 70 mg/DL in early life to 130 mg/dl at age 60 before slightly decreasing. Triglycerides observed another increase past 80 years of age. Mean LDL also observed a parabolic trend, increasing from 105 mg/dl at age 20 to 130 at age 60 before decreasing to 100 mg/DL at age 80.

### Vascular risk factor trajectories and CSVD score

The number of FPC scores retained for each risk factor ranged from three to five. We related the FPC scores to burden of CSVD where a higher score indicates higher CSVD burden. In ordinal logistic regression analyses, systolic blood pressure, diastolic blood pressure, pulse pressure, cigarettes per day, and triglycerides were all significantly associated with multi-marker CSVD score in the primary model (Table 3). The score test for the proportional odds assumption was not significant in any models and proportional odds were assumed. After adjusting for years of medication use, triglycerides were no longer significant (Supplementary Table S3).

**Table 3:** Ordinal logistic regression of Multi-marker CSVD scores on FPC scores for each risk factor adjusted for age at MRI, sex, and cohort.

| Risk Factor | OR (95% CI)<sup>†</sup> |  |  |  |  | Global p-value | Adjusted global p-value |
| --- | --- | --- | --- | --- | --- | --- | --- |
|  | PC1 | PC2 | PC3 | PC4 | PC5 |  |  |
| Systolic blood pressure, N=1625 | 1.22 (1.11, 1.35)<br>p < .0001 | 0.90 (0.79, 1.02)<br>p = 0.1131 | 0.92 (0.83, 1.01)<br>p = 0.0872 | 1.18 (1.06, 1.33)<br>p = 0.0047 | - | < 0.0001 | 0.0011 |
| Diastolic blood pressure, N=1625 | 1.11 (1.00, 1.22)<br>p = 0.0439 | 1.07 (0.94, 1.23)<br>p = 0.3304 | 0.96 (0.85, 1.07)<br>p = 0.4559 | 0.85 (0.75, 0.96)<br>p = 0.0154 | - | 0.0020 | 0.0073 |
| Pulse pressure, N=1671 | 1.21 (1.08, 1.35)<br>p = 0.0009 | 0.94 (0.84, 1.06)<br>p = 0.3334 | 0.92 (0.83, 1.01)<br>p = 0.0873 | 1.14 (1.02, 1.29)<br>p = 0.0290 | - | 0.0009 | 0.0049 |
| Cigarettes per day, N=1625 | 1.18 (1.07, 1.30)<br>p = 0.0011 | 1.01 (0.9, 1.12)<br>p = 0.9099 | 1.04 (0.94, 1.15)<br>p = 0.4656 | - | - | 0.0031 | 0.0085 |
| Body mass index, N=1617 | 0.94 (0.84, 1.06)<br>p = 0.3189 | 1.00 (0.88, 1.12)<br>p = 0.9400 | 1.05 (0.93, 1.18)<br>p = 0.4114 | 1.08 (0.96, 1.21)<br>p = 0.2021 | - | 0.4116 | 0.6468 |
| Fasting blood glucose*, N=1324 | 1.04 (0.92, 1.19)<br>p = 0.5201 | 1.08 (0.91, 1.29)<br>p = 0.3489 | 0.93 (0.77, 1.12)<br>p = 0.4542 | 0.96 (0.79, 1.17)<br>p = 0.6692 | - | 0.4945 | 0.6799 |
| Blood glucose*, N=142 | 1.12 (0.69, 1.83)<br>p = 0.6413 | 0.71 (0.34, 1.48)<br>p = 0.3486 | 0.91 (0.62, 1.34)<br>p = 0.6173 | 0.74 (0.43, 1.27)<br>p = 0.2675 | 0.57 (0.34, 0.96)<br>p = 0.0336 | 0.2771 | 0.5080 |
| Total cholesterol, N=1612 | 1.00 (0.90, 1.11)<br>p = 0.9867 | 0.98 (0.87, 1.1)<br>p = 0.7461 | 1.03 (0.92, 1.15)<br>p = 0.6337 | 0.96 (0.87, 1.06)<br>p = 0.4251 | - | 0.9478 | 0.9478 |
| High density lipoprotein, N=1596 | 0.92 (0.82, 1.04)<br>p = 0.1783 | 1.05 (0.90, 1.21)<br>p = 0.5498 | 0.94 (0.82, 1.06)<br>p = 0.3064 | 0.93 (0.81, 1.07)<br>p = 0.3245 | - | 0.6087 | 0.7440 |
| Low density lipoprotein, N=1458 | 1.02 (0.91, 1.14)<br>p = 0.7286 | 0.97 (0.86, 1.09)<br>p = 0.6062 | 0.94 (0.84, 1.05)<br>p = 0.2662 | 1.01 (0.89, 1.15)<br>p = 0.8889 | - | 0.7361 | 0.8097 |
| Triglycerides, N=1512 | 1.05 (0.92, 1.20)<br>p = 0.4390 | 1.16 (0.99, 1.36)<br>p = 0.0715 | 0.84 (0.72, 0.98)<br>p = 0.0335 | 0.92 (0.83, 1.03)<br>p = 0.1486 | - | 0.0199 | 0.0438 |

CI: confidence interval; OR = odds ratio; PC = principal component
\*Fasting blood glucose was measured in the Offspring cohort, random blood glucose in the Original cohort
† Odds ratios are for presented for a one standard deviation change in the principal component score

### Vascular risk factor trajectory clusters

Figure 4 illustrates the mean trajectories of the clusters for each risk factor.

**Figure 4:**
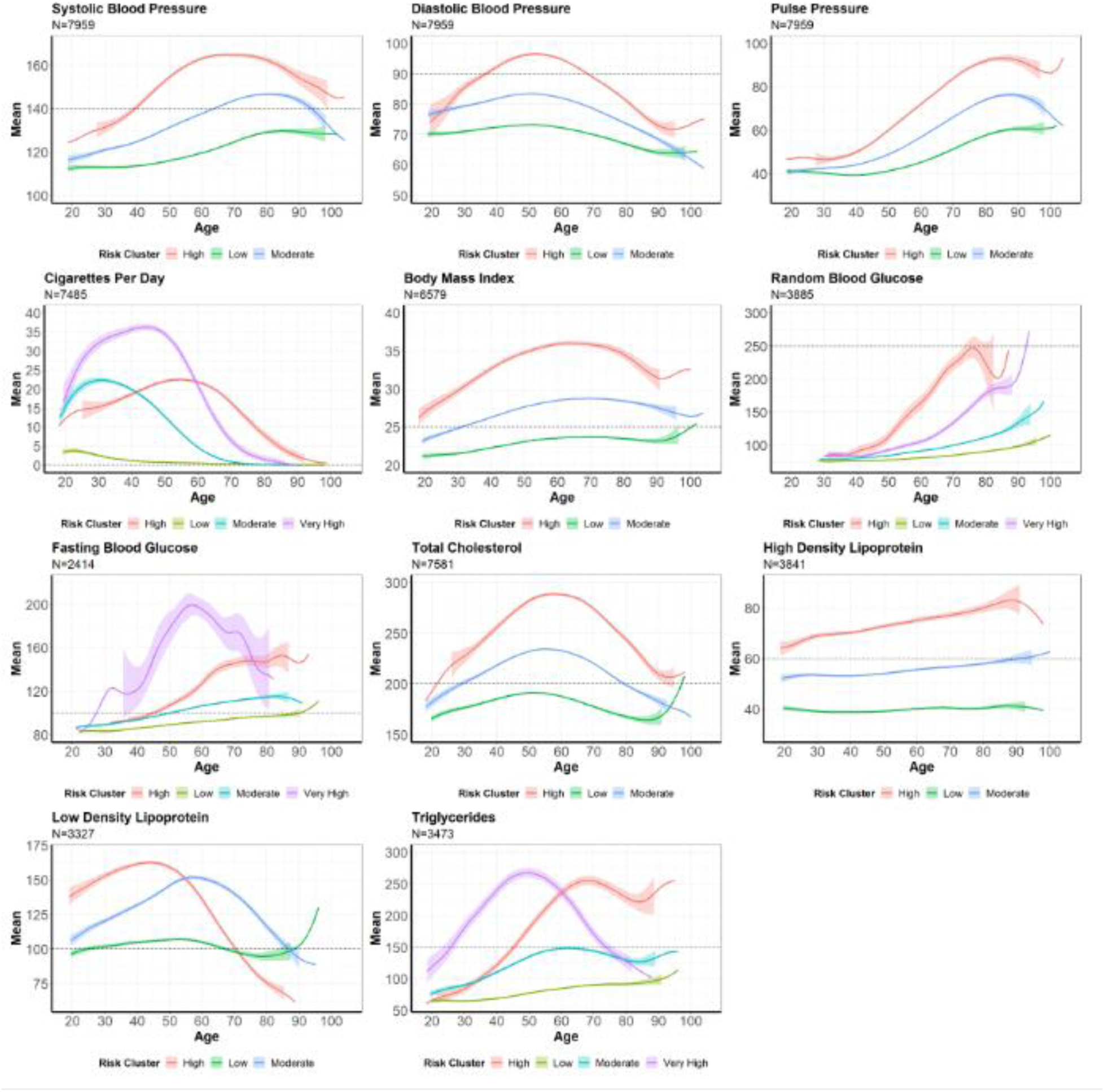
Mean Trajectories by Vascular Risk Factor Cluster Assignment.

Across multiple cardiometabolic risk factors, distinct life-course trajectory clusters revealed consistent separation between low-, moderate-, high-, and highest-risk groups. For systolic blood pressure (SBP), moderate- and high-risk clusters showed steep increases, exceeding 140 mmHg by ages 60 and 40 respectively, before peaking around ages 70–80 and then declining. The low-risk cluster rose gradually but remained below the hypertensive threshold across the lifespan. Diastolic blood pressure (DBP) patterns showed that only the high-risk cluster reached hypertensive levels (≥90 mmHg) between ages 40 and 70, peaking near age 50; in contrast, low- and moderate-risk clusters stayed below 90 mmHg, with moderate-risk individuals peaking at 85 mmHg around age 50.

For pulse pressure, moderate- and high-risk clusters were stable until age 40, after which the high-risk cluster increased sharply and the moderate-risk cluster rose gradually. Low-risk individuals remained near 40 mmHg until age 50 and then increased to about 60 in late life. Pulse pressure declined slightly in survivors beyond age 90 but never returned to levels seen in early life.

Smoking trajectories showed four clear patterns. The lowest-risk cluster engaged in only light smoking in early adulthood and quit by age 60. All other clusters exhibited decades of heavy use, with moderate- and high-risk groups peaking at similar intensities (22–23 cigarettes/day) but at different ages. The highest-risk cluster showed extremely heavy smoking, peaking at 35 cigarettes/day in the early 40s and remaining above 5 cigarettes/day even after age 70.

BMI clusters all increased with age but diverged in magnitude. The high-risk cluster maintained BMI >25 kg/m² throughout life, exceeded the obesity threshold at age 30, and peaked at 35 kg/m² around age 60 before declining slightly but remaining obese. The moderate-risk cluster became overweight by age 30 and peaked around age 70 without entering the obese range. Low-risk individuals sustained normal BMI levels across the lifespan.

Patterns for glucose differed between the Original and Offspring cohorts. In the Original cohort (random glucose), all clusters remained below 100 mg/dL before age 45; later-life peaks reached 110 mg/dL in the low-risk group, 160 mg/dL in the moderate-risk group, and ≥250 mg/dL in the two highest-risk clusters. In the Offspring cohort (fasting glucose), all groups began below 100 mg/dL; the moderate and high-risk clusters exceeded 100 mg/dL by midlife, with the high-risk cluster rising toward 140 mg/dL in late life, while the highest-risk cluster surged to 200 mg/dL by age 55 before decreasing to 130 mg/dL at age 80.

For total cholesterol, the high-risk cluster exceeded 200 mg/dL from age 20 onward, peaking around age 60. The moderate-risk cluster crossed 200 mg/dL by age 30 and peaked near 230 mg/dL before returning to normal levels by age 80. Low-risk individuals stayed mostly within normal range, with only a slight late-life rise above threshold.

Across lipid subtypes, patterns varied. HDL levels increased steadily for low- and moderate-risk clusters, both reaching >60 mg/dL in late life; the high-risk cluster maintained consistently low levels near 40 mg/dL. LDL levels in the high-risk group were elevated early, peaking at 160 mg/dL before declining to optimal levels by age 70. The moderate-risk cluster exceeded 100 mg/dL throughout life and peaked at 150 mg/dL around age 60. Low-risk individuals stayed near 100 mg/dL until a rise after age 90. Triglyceride trajectories showed graded severity: low-risk individuals increased gradually to 100 mg/dL, moderate-risk individuals peaked near 150 mg/dL around age 60, and high- and highest-risk clusters reached 250–260 mg/dL before late-life declines.

### Vascular risk factor trajectory clusters and CSVD score

There were significant global associations between CSVD score and risk factor clusters for systolic blood pressure and pulse pressure; cigarettes per day did not meet the FDR-adjusted significance threshold but showed a borderline association (adjusted p = 0.0513) (Table 4). Individuals in the moderate risk cluster for SBP had 39% increased odds of being in a higher score category compared to the low-risk cluster (OR: 1.39, 95% CI: 1.14-1.69) while those in the high-risk cluster had 70% increased odds (OR: 1.70; 95% CI: 0.97-3.00). Similar odds ratios were observed for pulse pressure.

**Table 4:** Ordinal logistic regression of multi-marker CSVD scores on cluster assignments for each risk factor adjusted for age at MRI, sex, and cohort.

|  | Low Risk Cluster |  | Moderate Risk Cluster |  | High Risk Cluster |  | Very High Risk Cluster |  |  |  |
| --- | --- | --- | --- | --- | --- | --- | --- | --- | --- | --- |
| Risk Factor | n | OR (95% CI) | n | OR (95% CI) | n | OR (95% CI) | n | OR (95% CI) | Global p-value | Adjusted global p-value |
| Systolic blood pressure, N=1625 | 981 | REF | 599 | 1.39 (1.14, 1.69)<br>p = 0.0011 | 45 | 1.70 (0.97, 3.00)<br>p = 0.0650 |  | - | 0.0018 | 0.0176 |
| Diastolic blood pressure, N=1625 | 878 | REF | 688 | 1.10 (0.90, 1.34)<br>p = 0.3603 | 59 | 1.10 (0.66, 1.86)<br>p = 0.7090 |  | - | 0.6470 | 0.7845 |
| Pulse pressure, N=1671 | 921 | REF | 614 | 1.32 (1.08, 1.61)<br>p = 0.0061 | 90 | 1.70 (1.12, 2.58)<br>p = 0.0124 |  | - | 0.0032 | 0.0176 |
| Cigarettes per day, N=1625 | 1244 | REF | 266 | 1.48 (1.15, 1.91)<br>p = 0.0025 | 55 | 1.39 (0.83, 2.33)<br>p = 0.2118 | 60 | 1.30 (0.79, 2.15)<br>p = 0.3034 | 0.0140 | 0.0513 |
| Body mass index, N=1617 | 666 | REF | 745 | 0.76 (0.62, 0.93)<br>p = 0.0094 | 206 | 0.79 (0.58, 1.08)<br>p = 0.1335 |  | - | 0.0285 | 0.0784 |
| Fasting blood glucose*, N=1324 | 796 | REF | 416 | 1.06 (0.83, 1.35)<br>p = 0.6523 | 81 | 0.85 (0.54, 1.33)<br>p = 0.4776 | 31 | 2.46 (1.24, 4.86)<br>p = 0.0100 | 0.0572 | 0.1258 |
| Blood glucose*, N=142 | 121 | REF | 15 | 0.90 (0.32, 2.51)<br>p = 0.8441 | - | - | 6 | 1.09 (0.23, 5.23)<br>p = 0.9189 | 0.9739 | 0.9739 |
| Total cholesterol, N=1612 | 909 | REF | 630 | 0.94 (0.77, 1.16)<br>p = 0.5801 | 73 | 1.12 (0.69, 1.82)<br>p = 0.6534 |  | - | 0.7132 | 0.7845 |
| High density lipoprotein, N=1596 | 351 | REF | 625 | 1.05 (0.81, 1.36)<br>p = 0.7227 | 620 | 1.18 (0.89, 1.57)<br>p = 0.2590 |  | - | 0.4655 | 0.7315 |
| Low density lipoprotein, N=1458 | 705 | REF | 367 | 0.91 (0.71, 1.17)<br>p = 0.4654 | 368 | 1.05 (0.82, 1.35)<br>p = 0.6851 |  | - | 0.5935 | 0.7845 |
| Triglycerides, N=1512 | 782 | REF | 520 | 1.09 (0.88, 1.36)<br>0.4284 | 103 | 1.61 (1.09, 2.38)<br>0.0170 | 107 | 1.24 (0.83, 1.85)<br>0.2908 | 0.0976 | 0.1789 |

CI: confidence interval; OR = odds ratio; PC = principal component
\*Fasting blood glucose was measured in the Offspring cohort, random blood glucose in the Original cohort

Although the global BMI–CSVD association was not significant, the moderate-risk BMI cluster had lower odds of higher CSVD scores than the low-risk cluster (OR = 0.76, 95% CI: 0.62–0.93), while the high-risk cluster’s reduction was not significant (OR = 0.79, 95% CI: 0.58–1.08). Similarly, the moderate, high, and very high-risk clusters for cigarettes smoked per day observed increased odds of having higher CSVD scores compared to those in the low-risk cluster. However, only the moderate risk cluster observed an increase in odds (OR: 95% CI: 1.48, 95% CI: 1.15-1.91).

After additionally adjusting for years of medication use, the associations of CSVD score with clusters for systolic blood pressure, pulse pressure, cigarettes per day, and BMI remained significant (Supplementary Table S4).

## Discussion

We evaluated the lifelong trajectories of common modifiable vascular risk factors in community-dwelling participants of the Framingham Heart Study to explore their associations with markers of advanced cerebral small vessel disease. Lifetime trajectories of systolic and diastolic blood pressure, pulse pressure, number of cigarettes per day, and triglycerides were significantly associated with the multi-marker CSVD score. In addition, we identified three to four clusters of trajectories for each vascular risk factor to detect groups at heightened risk for advanced CSVD and these clusters were separated at the onset of the trajectory suggesting that this separation may occur even at earlier stages during childhood. Cluster assignment for systolic blood pressure and pulse pressure were significantly associated with the multi-marker CSVD score and borderline association was observed with cigarettes per day.

In contrast to other studies that assessed longitudinal risk factors in relation to various outcomes, our study is unique in the characterization of life-long trajectories of VRF spanning up to seven decades of follow up. Our observations expand current knowledge regarding trajectories in risk factors over a lifetime in a large sample of community dwelling individuals and provide insight that may inform further preventive efforts, public health policy and, through the assessment of the risk factor trajectory, identify potential subgroups of individuals with under-recognized increased vascular risk. Prior studies have been limited to assessing single risk factors, or multiple risk factors over a much shorter period, and have not assessed the relation to burden of CSVD.^9–11^

In our sample, mean SBP levels declined later in life, possibly reflecting the impact of public health efforts on hypertension awareness and the increased use of antihypertensive medication, but other physiological factors may contribute. Additionally, SBP trends and cluster assignment were both significantly associated with CSVD score, suggesting that not only individual variations in SBP patterns but also broader trajectory classifications are important in relation to CSVD burden.

In general, blood pressure patterns reflect the typical progression of hypertension over the course of life, emphasizing the prolonged exposure many individuals experience. The complementary insights from systolic, diastolic, and pulse pressure measures further illustrate these trends. Given that hypertension is one of the most prevalent risk factors in the population, affecting nearly 50% of Americans,^12^ our observation is highly relevant to inform public health efforts. Public education of the risks of high blood pressure should be initiated early in life and actively maintained throughout adult life. Such educational efforts should include informing of the adverse cerebrovascular effects of hypertension as noted by the association with advanced CSVD in our study. Pulse pressure also showed patterns that likely reflect an increase in central arterial stiffness around the fifth decade of life and advancing thereafter, likely affecting cerebral vascular beds and clinical outcomes, as shown by cluster analysis.

In our study, the decline in mean cigarettes per day over the lifespan likely reflects the impact of public health efforts to reduce smoking. Overall trends in cigarette smoking and cluster assignment were significantly associated with CSVD burden, underscoring its status as a major public health concern. This observation brings attention to the need of continued public health education and regulatory efforts to minimize smoking in the general population.

We observed that overall BMI trends were not significantly associated with CSVD score. However, specific groups of individuals with distinct BMI trajectories showed an association, suggesting that certain patterns of BMI change, rather than overall variation, may be more relevant to CSVD burden in late life. Notably, the relation of BMI and CSVD burden showed an inverse pattern where clusters representing trends with longest exposure to overweight and obesity were associated with lower risk of advanced CSVD. Low BMI has been suggested as a marker of frailty and sarcopenia, which have also been related to markers of CSVD such as white matter hyperintensities.^13^ However, residual confounding in these associations by other factors such as lifestyle or other comorbidities is plausible.

Despite evidence of uncontrolled blood glucose in our sample, neither trajectories nor clusters for random or fasting blood glucose reached the statistical significance threshold. While elevated blood glucose is a known risk factor for vascular disease, its relationship with CSVD burden may be influenced by concurrent risk factors and their treatment effects. Despite an elevated risk of having high CSVD scores, the highest risk cluster for random blood glucose did not reach statistical significance. This group showed greatly elevated blood glucose levels throughout the life course and is likely an under-recognized target for cardiovascular prevention.

Mean trajectories for total cholesterol and LDL were observed to increase from early to mid-life before steadily declining, likely reflecting the stage where individuals began lipid lowering therapies. For HDL, overall and by cluster, mean levels gradually increased over time. Total cholesterol, HDL, and LDL were not significantly associated with CSVD burden despite being known risk factors for cardiovascular and cerebrovascular disease. It is possible that the lipid levels attained in our sample, which were lower than those typically observed in clinical samples, may not reach the threshold for an association with CSVD burden, but these findings don’t exclude an association in individuals not represented in our sample.

In general, triglycerides trends increased from early to mid-life and only declined in higher risk clusters, remaining suboptimal for nearly four decades. Individual trends of triglycerides were significantly associated with CSVD burden. This highlights triglycerides as an important modifiable risk factor where early treatment may reduce risk of CSVD. However, despite increased risk of higher CSVD scores, there was no global association of triglyceride cluster with CSVD score suggesting that there may be other metabolic or vascular factors not fully accounted for by cluster-based classification.

Given that CSVD is the most frequent underlying type of cerebrovascular disease in patients with stroke, dementia and cognitive disorders, disproportionately affecting populations in developing countries, our findings highlight the importance of recognizing the trajectories of modifiable risk factors. Such recognition is essential for creating preventive strategies to mitigate their impact on cerebral circulation and overall vascular health. In addition, our findings suggest that use of current vascular risk factor treatments alone may not be sufficient to ameliorate CSVD burden. Effective management of VRF likely requires ongoing monitoring to ensure treatment targets are sustained over time, in combination with the development and implementation of novel treatments aimed at targeting key aspects in CSVD pathophysiology. Our observations are in line with a recent scientific statement from the American Heart Association highlighting the importance of covert cerebrovascular disease recognition for the prevention of vascular cognitive impairment and dementia,^14^ with major implications for public health preventive interventions that could benefit millions of Americans.

Our study has many strengths including the prospective cohort design, inclusion of a large sample of community dwelling participants closely followed over time comprising the longest duration of follow-up of any epidemiological study, and blinded assessment of exposures and CSVD outcomes with high reliability. However, several limitations also need consideration. While long-term follow-up data were available, detailed assessments between exam cycles were not available, thus not capturing granular variability in risk factor levels over time, particularly in high-risk groups. However, this type of assessment is what is followed in clinical practice where intermittent assessments of risk factors guide clinical care. Additionally, cohort differences in exam frequency may affect trajectory estimation: Gen1 participants typically contribute more longitudinal measurements than Gen2, potentially yielding more stable FPCA scores. Age-based alignment and adjusting for cohort in regression models aims to mitigate these differences.

Our sample is of predominantly White racial background thus limiting generalization to other racial groups. The sample of participants excluded from the trajectories of vascular risk factors analyses had a higher vascular risk at baseline (Table 1) than those included, which may have led to an underestimation of risk in our findings, as those with the highest vascular burden were not fully represented. On the other hand, the sample based on MRI availability showed similar baseline characteristics among included and excluded participants.

Finally, survivor bias is likely present as participants with the highest vascular risk may have been more likely to experience adverse health outcomes, including death or dropout, leading to reduced data availability in late life, as evidenced by the wide confidence bands at these ages in mean trajectories. This could result in an underestimation of the true burden of VRFs on CSVD, as those who survived and remained in the study may represent a healthier subset of those who were originally enrolled. To address survivor bias, future analyses could formally address informative dropout using inverse-probability weights for survival and MRI participation, and age-restricted models (e.g., truncating at ≤80 years).

## Conclusions

This work used lifelong vascular risk factor trajectory data in a large sample of community dwelling individuals observing that individuals cluster in three to four risk groups, which vary in age at onset and duration of exposure to abnormal levels for individual vascular risk factors. In turn, the trajectory risk groups relate in a dose-effect manner to burden of CSVD. Our findings support consideration of an individual’s trajectory in risk factors for preventive efforts to minimize vascular brain injury, and potentially its adverse consequences including stroke and dementia. Our findings also bring attention to targets for public health interventions to further impact the treatment of modifiable vascular risk factors at the population level, which may result in major prevention of major cardiovascular events and dementia.

## Data Availability

Requests to access de-identified data from the Framingham Heart Study can be made at https://www.framinghamheartstudy.org/fhs-for-researchers.

## References

1. de Havenon A, Sheth KN. Stroke’s Resurgence as a Leading Cause of Death in the United States. Stroke. 2025;56:e199–e200. doi: 10.1161/STROKEAHA.125.052010

2. Joynt Maddox KE, Elkind MSV, Aparicio HJ, Commodore-Mensah Y, de Ferranti SD, Dowd WN, Hernandez AF, Khavjou O, Michos ED, Palaniappan L, et al. Forecasting the Burden of Cardiovascular Disease and Stroke in the United States Through 2050-Prevalence of Risk Factors and Disease: A Presidential Advisory From the American Heart Association. Circulation. 2024;150:e65–e88. doi: 10.1161/CIR.0000000000001256

3. Collaborators GBDDF. Estimation of the global prevalence of dementia in 2019 and forecasted prevalence in 2050: an analysis for the Global Burden of Disease Study 2019. Lancet Public Health. 2022;7:e105–e125. doi: 10.1016/S2468-2667(21)00249-8

4. Lara FR, Scruton AL, Pinheiro A, Demissie S, Parva P, Charidimou A, Francis M, Himali JJ, DeCarli C, Beiser A, et al. Aging, prevalence and risk factors of MRI-visible enlarged perivascular spaces. Aging (Albany NY). 2022;14:6844–6858. doi: 10.18632/aging.204181

5. Duering M, Biessels GJ, Brodtmann A, Chen C, Cordonnier C, de Leeuw FE, Debette S, Frayne R, Jouvent E, Rost NS, et al. Neuroimaging standards for research into small vessel disease-advances since 2013. Lancet Neurol. 2023;22:602–618. doi: 10.1016/S1474-4422(23)00131-X

6. Pinheiro A, Ekenze O, Aparicio HJ, Beiser AS, Decarli CS, Demissie S, Seshadri S, Romero JR. Multimarker Cerebral Small Vessel Disease Score and Risk of Incident Dementia in the Framingham Heart Study. Neurology. 2025;105:e214113. doi: 10.1212/WNL.0000000000214113

7. Yao F MuH, and Wang JL. Functional data analysis for sparse longitudinal data. J Am Stat Assoc. 2005;100:577–590.

8. Pinheiro A, Aparicio H, Lioutas VA, Beiser A, Ekenze O, DeCarli C, Seshadri S, Demissie S, Romero JR. Higher Burden of Cerebral Small Vessel Disease Is Associated With Risk of Incident Stroke in Community-Dwelling Individuals. J Am Heart Assoc. 2025;14:e040263. doi: 10.1161/JAHA.124.040263

9. Sedaghat S, Lutsey PL, Ji Y, Empana JP, Sorond F, Hughes TM, Mosley TH, Gottesman RF, Knopman DS, Walker KA, et al. Association of change in cardiovascular risk factors with incident dementia. Alzheimers Dement. 2023;19:1821–1831. doi: 10.1002/alz.12818

10. Liang Y, Ngandu T, Laatikainen T, Soininen H, Tuomilehto J, Kivipelto M, Qiu C. Cardiovascular health metrics from mid- to late-life and risk of dementia: A population-based cohort study in Finland. PLoS Med. 2020;17:e1003474. doi: 10.1371/journal.pmed.1003474

11. Jansen MG, Griffanti L, Mackay CE, Anaturk M, Melazzini L, Lange AG, Filippini N, Zsoldos E, Wiegertjes K, Leeuw FE, et al. Association of cerebral small vessel disease burden with brain structure and cognitive and vascular risk trajectories in mid-to-late life. J Cereb Blood Flow Metab. 2022;42:600–612. doi: 10.1177/0271678X211048411

12. Martin SS, Aday AW, Almarzooq ZI, Anderson CAM, Arora P, Avery CL, Baker-Smith CM, Barone Gibbs B, Beaton AZ, Boehme AK, et al. 2024 Heart Disease and Stroke Statistics: A Report of US and Global Data From the American Heart Association. Circulation. 2024;149:e347–e913. doi: 10.1161/CIR.0000000000001209

13. Kant IMJ, Mutsaerts H, van Montfort SJT, Jaarsma-Coes MG, Witkamp TD, Winterer G, Spies CD, Hendrikse J, Slooter AJC, de Bresser J, et al. The association between frailty and MRI features of cerebral small vessel disease. Sci Rep. 2019;9:11343. doi: 10.1038/s41598-019-47731-2

14. Smith EE, Aparicio HJ, Gottesman RF, Goyal MS, Greenberg SM, Schneider JA, Sorond FA, Wright CB, American Heart Association Stroke C, Council on C, et al. Vascular Contributions to Cognitive Impairment and Dementia in the United States: Prevalence and Incidence: A Scientific Statement From the American Heart Association. Stroke. 2025;56:e317–e330. doi: 10.1161/STR.0000000000000494

